# Sexually Transmitted Infections Among Patients on Antiretroviral Treatment at Adama Hospital Medical College, Adama, Ethiopia

**DOI:** 10.1101/2024.01.03.24300757

**Authors:** Midekso Sento, Atoma Negera, Samuel Negera

## Abstract

**Background:** Sexually transmitted infections (STIs) are an infections or clinical syndromes caused by microorganisms that can be acquired and transmitted via sexual contact. Sexually transmitted infections are a major public health problem worldwide that cause acute illness, long-term complications, infertility, medical as well as psychological consequences, and death. Moreover, it facilitates the spread of the human immunodeficiency virus (HIV). To develop policies and strategies targeted at preventing and controlling illnesses, research on sexually transmitted infections in general and among people with HIV in particular is crucial.

**Objective:** to determine the prevalence and associated factors of sexually transmitted infections among human immunodeficiency virus patients in antiretroviral treatment clinic in Adama Hospital Medical College (AHMC), Adama, Ethiopia, 2022.

**Methods:** an institution-based, retrospective cross-sectional study design among 376 HIV patients in Anti-retroviral treatment clinic in AHMC was conducted on January-February, 2022. Study participants were selected using a systematic sampling. Data was collected using a structured questionnaire and was taken from individual cards with a prepared checklist. The collected data was processed and analyzed with Statistical Package for Social Science (SPSS) version 25.0. Descriptive statistical analyses were done to define the characteristics of the participants. Variables with p-value ≤0.25 in the bivariable analyses were entered into the multivariable logistic regression analysis to identify factors associated with STIs. Adjusted Odds Ratio (aOR) with a 95% Confidence Interval (CI) was computed and variable with a p-value less than 0.05 was declared as statistically significant variable.

**Results:** among 376 HIV positive patients 39 (10.37%) have STI and among them, 66.7% were female patients. Those under 35 years had 3.03 times more STI (aOR=3.03; 95% CI: (1.3, 6.66)) than those above 35 years. Those with CD4 of <500 had 11.7 times (aOR-=11.7; 95% C: (4.6, 30)) more STI than those with CD4 of > 500. Those with multiple sexual partners had 9.2 times (aOR=9.2; 95% CI: (2.7, 30.8)) more STI than those without multiple partners.

**Conclusion:** the study revealed among 376 HIV-positive patients 39(10.37%) have STI and 337 (89.63%) have no STI. Age (lower age group), having multiple sexual partners, and lower CD4 count were the factors significantly associated with STIs. Thus, we recommend health workers should take time in counseling ART patients about STI in every visit.

## 1. INTRODUCTION

Sexual Transmitted Infections (STIs) are a group of clinical disorders caused by pathogens that can be acquired and transmitted through sexual interaction. More than thirty viral, bacterial, and parasite infections are transmitted sexually [1]. STIs are a significant global public health issue that can result in acute illness, long-term complications, infertility, negative physical and psychological effects, and even death [2]. Moreover, it facilitates the spread of the human immunodeficiency virus (HIV) [3]. In 2012, there were 498.9 and 92.6 million new STI cases worldwide and in Africa, respectively. Consequently, on average 1.4 million people worldwide contract an STI every day [4]. In Ethiopia, those aged 15 to 24 have the highest reported incidence of STIs, and they also account for 60% of all new HIV infections and roughly 50% of all people with HIV [5]. A person who has an untreated STI, particularly one that results in discharge or ulcers, is more likely to transmit or get HIV during sexual activity. When an STI is present, there is a higher chance that the virus will enter or leave the body through damaged skin or mucous membranes. Since the virus is attempting to infect these cells, the cells battling the infection will concentrate there. Recent research indicates that vaginal ulcers caused by syphilis, chancroids, or genital herpes (HSV-2) multiply the chance of developing HIV by a factor of 10 to 300 after a single exposure [6]. Recent studies, using molecular techniques for detecting viral RNA, have shown that STI clinic attendees in the US [7], Malawi [8] and India [9] can have a surprisingly high prevalence of acute HIV infection.

In 1991, patients treated for various STIs had an HIV seroprevalence rate of 12% [10], but by 2001, patients with genital ulcer disorders had an HIV seroprevalence rate of 64% [23]. Additionally, with HIV seroprevalence rising from 16%, a considerable increase in the relationship between genital ulcer disorders and HIV has been observed [10] to 90% [11] among individuals with genital ulcer diseases. According to other recent studies carried out in Ethiopia, HSV-2 seroprevalence is substantially greater in HIV-positive persons (61.5%) than it is in HIV-uninfected people (29.1%) [12] and can reach 87% in patients with STIs [13]. The majority of the world’s regions still consider STIs to be a severe public health concern. HIV and STI convection interact in both directions. This is because STIs play a crucial role in HIV acquisition and transmission and both HIV and common STIs affect STI treatment and predispose to HIV infection [15]. The prevalence of numerous STDs among HIV-positive patients is significantly higher than it is among HIV-negative controls in the majority of areas of the world [16].

Since the immune system is more weakened in HIV-positive patients than in HIV-negative controls, the prognosis is significantly worse for numerous STDs. For example, syphilis and HIV may manifest differently and have a propensity to develop neurosyphilis fast. Atypical lesions, which can be big, numerous, and accompanied by fever and chills, are frequently developed by chancroids [14]. Adolescence is a time for experimentation and taking chances, frequently without considering the possible repercussions. In addition, most people have their first sexual interactions during this period, whether or not they end up getting married [17]. Ninety percent of STD cases recorded globally occur in underdeveloped countries, and of those, 25% are in people under the age of 20. The presence of STIs increases the risk of STIs, including HIV, because it indicates that they engaged in unprotected sexual activity [18]. HIV Outpatient Programme in New Orleans began receiving routine Chlamydia and gonorrhea testing from HIV/STD programs in Louisiana. Although there was no discernible difference in STD prevalence by sex or race, it was higher in those under the age of 30 [19]. The London syphilis outbreak, which is the largest in the UK to date and shows the burden of new diagnoses among men who have sex with men and many of whom are HIV-positive, is a major public health concern [20]. In low resource countries, STIs including HIV is a major public health concern which needs attention of different organization and comprehensive study to alleviate the burden. Therefore, this study has two aims: the first one is to assess prevalence of STIs among patients on ART. Secondly, to identify factors associated with STIs among patients on ART.

## METHODS

### Ethical consideration

Ethical clearance was obtained from the ethical review committee of AHMC. Due to the nature of the study, consent was not obtained directly from the patients. However, an official letter was submitted to AHMC and permission was obtained. The purpose of the study was explained to each worker to precede the data collection from patient registration. Information that could identify individual participants was not collected for analysis.

### Study Design

The study was conducted using a facility-based retrospective cross-sectional study employed from patient charts from January 01 to February 28, 2022 to assess magnitude of STI among HIV-positive patients who were linked to ART clinic at Adama Hospital and Medical College from September 2020 to September 2021.

### Study area

The study was conducted in Adama Hospital and Medical College, Adama town, Oromia region, Ethiopia. Adama is located at 99 km southeast of Addis Ababa (capital city of Ethiopia). Adama Hospital and Medical College is one of Ethiopia’s oldest hospital established in 1946 by missionaries from overseas. Subsequently, the hospital came under government control in 1978. The hospital has 1200 beds and serves more than 5 million people from the eastern part of the country.

### Participants

All HIV-positive patients who were linked to ART clinic at Adama Hospital and Medical College from September 2020 to September 2021 were extracted from the data base. All complete clinical records registered between September 2020 to September 2021 were eligible for the study. Any patient chart with missing clinical records or incomplete data were excluded.

### Sample size determination

The required sample size was estimated using the proportion of sexually transmitted infections based on the syndromic approach among HIV Patients in ART Clinic 42% which is taken from a retrospective institutional-based study done in Ayder Referral Hospital [21] with the assumption of margin of sampling error tolerated (d) 5%, and 95% confidence level.

n = ((Z_α/2_)^2^ *p*q)/d^2^ Where:

n= the desired sample size

Z_(α/2)_ =the critical value at a 95% level of significance (1.96) p=proportion of patients with STI, which is 0.42 d=precision of measurement (acceptable marginal error)

n= ((1.96)^2^*(0.42)*(0.58))/(0.05)^2^ = 372

In the AHMC ART clinic from September 2020 to September 2021, 4200 patients were linked to the clinic which makes the source population less than 10000 so the correction formula was used

Correction formula = n/1+(n/N) = 372/1+(372/4200) =341.9

After the addition of 10% for missed clinical records or incomplete data total sample size is 376.

### Sampling procedure

We used a systematic sampling method to select the required study participants. The interval was determined as k= N/n = 4200/376=11. Study subjects were selected every 11 intervals.

### Data collection

Data was collected from patient clinical records from January 01 to February 28, 2022 using a structured data collection format and a predefined checklist. The data collection checklist consists of 20 questions. It assesses the respondents’ socio-demographic characteristics, their sexual practice, and clinical characteristics. Before the initiation of data collection, five nurses who work in the ART clinic were trained and during the collection time, they were being supervised by the investigators.

### Data analysis

The collected data was cross checked for completeness and consistency. Then, entered and analyzed with SPSS 25.0 statistical software. Descriptive statistical analyses were done to define the characteristics of the participants. Variables with a p-value ≤0.25 in the bivariable analyses were entered into the multivariable logistic regression analysis to identify factors associated with STIs. Multicollinearity and model fitness were checked by the Variance inflation factor (VIF) and Hosmer and Lemeshow goodness-of-fit tests. None of the variables exhibited variance inflation factors greater than 10. The Hosmer and Lemeshow’s test produced an insignificant result (p-value = 0.578), indicating the suitability of the fitted model. The Adjusted Odds Ratio (aOR) with a 95% Confidence Interval (CI) was computed and variable with a p-value less than 0.05 was declared as statistically significant variable. The output data was presented as tables, and figures.

### Data Quality Assurance

The tool used for data collection was a structured questionnaire prepared in English. The questionnaire was pretested on 19 patients at a different setup (Hawas Health Center) and necessary modifications were made. Two hours training was given for data collectors before data collection. Regular supervision was done to check the consistency and completeness of the filled-out checklists, by the principal investigators.

## RESULTS

### Socio-demographic characteristics of the participants

From the total of 4200 HIV-positive patients linked to the ART clinic at AHMC 376 HIV-positive patients were selected by systematic sampling in this study. About two-third (66.48%) of the participants age group were 35 years and above. More than half (58.24%) respondents were female and 141 (37.5%) were married followed by single 121 (32.2%). The majority 108 (28.72%) of respondents’ occupational status were merchant followed by daily laborer 90 (23.9%) (Table 1). Among 376 HIV-positive patients, thirty-nine (10.37%) have STI, and three-hundred thirty-seven (89.63%) have no STI (Figure 1).

**Table 1.** Socio-demographic characteristics of HIV-positive patients linked to the ART clinic.

| Variables | characteristics | Frequency | % |
| --- | --- | --- | --- |
| Age | 15-24 | 37 | 9.8% |
|  | 25-34 | 88 | 23.4% |
|  | 35-44 | 155 | 41.22% |
|  | 45+ | 96 | 25.53% |
| Sex | Male | 157 | 41.76% |
|  | Female | 219 | 58.24% |
| Marital status | Single | 121 | 32.18% |
|  | Married | 141 | 37.5% |
|  | Divorced | 84 | 22.34% |
|  | Widowed | 30 | 7.97% |
| Occupation | Government Employee | 76 | 20.21% |
|  | NGO | 26 | 6.91% |
|  | House wife | 44 | 11.70% |
|  | Student | 23 | 6.11% |
|  | Merchant | 108 | 28.72% |
|  | Daily laborer | 90 | 23.93% |
|  | Farmers | 9 | 2.39% |

NGO- Non-Government Organization

**Figure 1.**
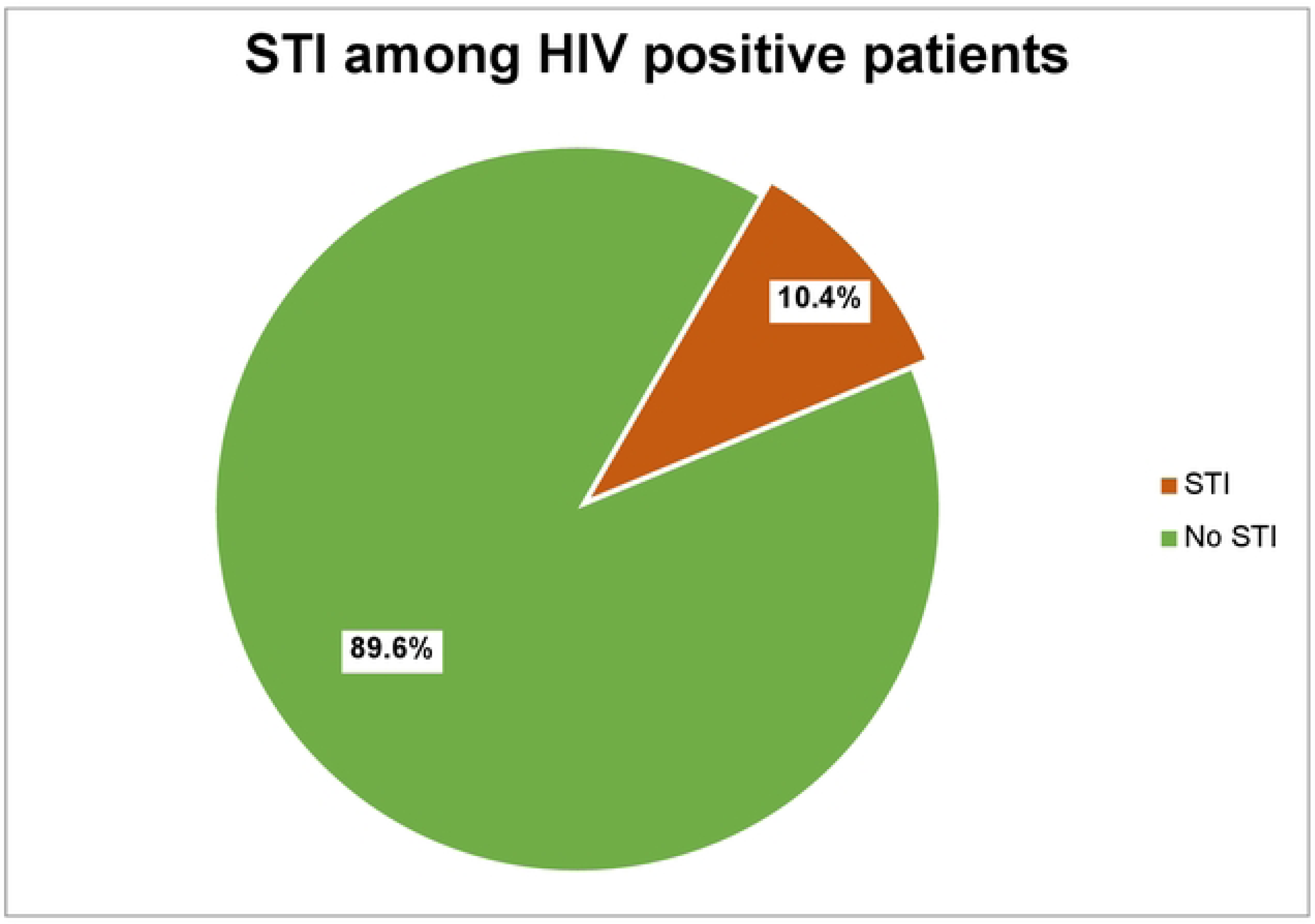
STI among HIV positive patients linked to ART clinic at AHMC.

### Distribution of common STI by age group

Among a total of 39 STI cases, vaginal discharge 5 (12.82%), genital ulcers 4(10.25%), and urethral discharge 3(7.69) are common in the 25-34 years of age group (Figure 2) (Table 2).

**Figure 2:**
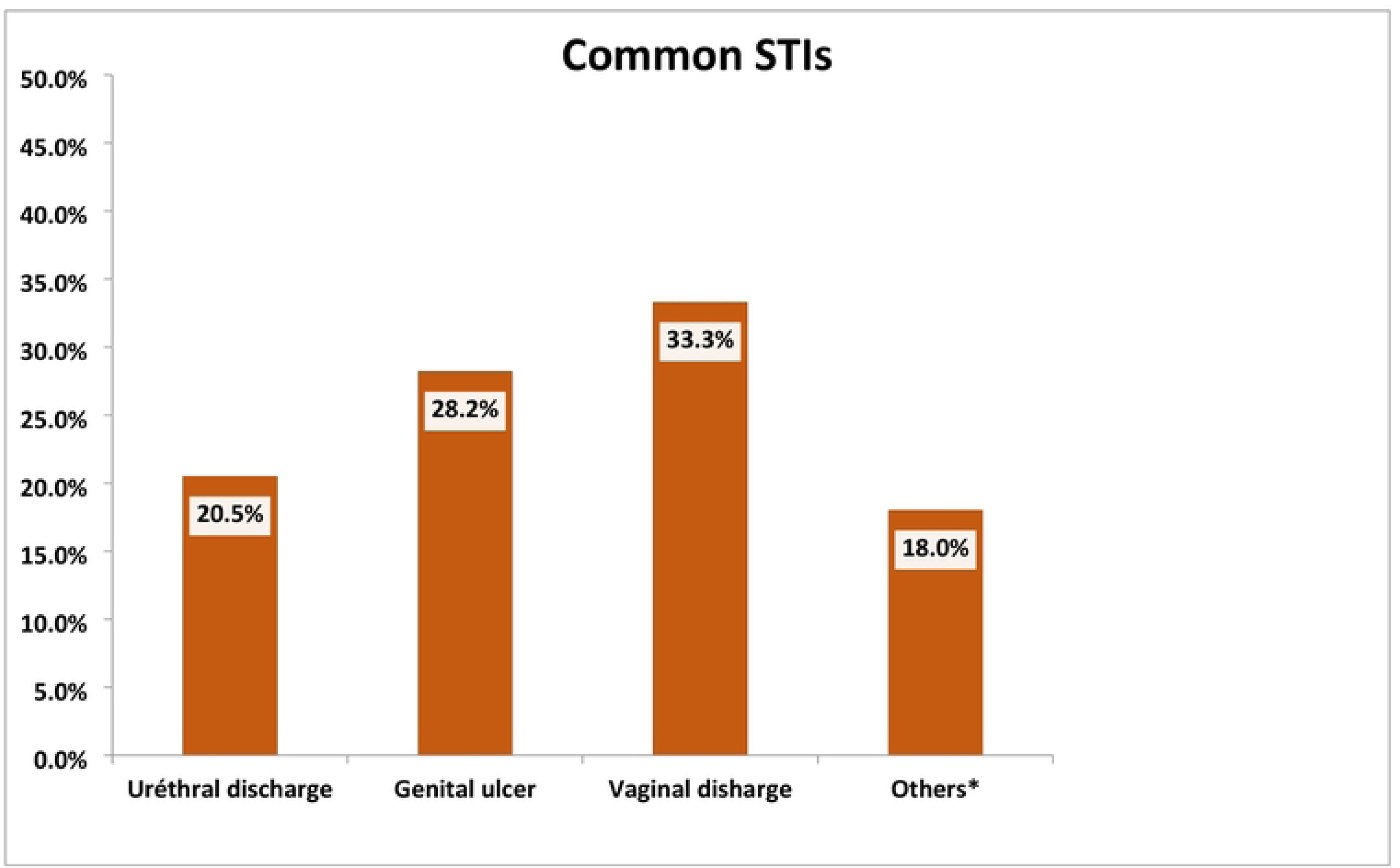
Three Common STIs among HIV-positive patients linked to the ART clinic at AHMC

**Table 2.** Distribution of common STI by age group among HIV-positive patients linked to ART clinic at AHMC.

| Age groups | Types of STI based on syndromic approach |  |  |  |  |  |
| --- | --- | --- | --- | --- | --- | --- |
|  | Urethral Discharge | Genital ulcer | Vaginal Discharge | Lower abdominal pain | Scrotal swelling | Inguinal bubo |
|  | No (%) | No (%) | No (%) | No (%) |  | No (%) |
| 15-24 | 2(5.12) | 3(7.69) | 2(5.12) | - | - |  |
| 25-34 | 3(7.69) | 4(10.25) | 5(12.82) | 3(7.69) | - |  |
| 35-45 | 3(7.69) | 3(7.69) | 4(10.25) | 2(5.12) | 1(2.56) |  |
| 45+ | - | 1(2.56) | 2(5.12) | - | 1(2.56) |  |
| Total | 8(20.5) | 11(28.19) | 13(33.31) | 5(12.81) | 2(5.12) |  |

### Personal Habit and Practice of Study Subjects

Thirty (7.97%) respondents had a new sexual partner within the last three months, of them, 22 (73.3%) respondents had one new sexual partner, and the rest 26.7% had more than one new sexual partner and 68.1% of the participants used condom regularly during sexual intercourse.

### Disease-Related Clinical Profiles of Study Subjects

Thirteen male and twenty-six female patients were diagnosed with STI, of them 8 patients (20.5%) had urethral discharge syndrome, 11 (28.2%) had genital ulcer syndrome, 2 (5.12) scrotal swelling syndrome, 5 (12.8%) had lower abdominal pain syndrome, and 13 (33.3%) patients had vaginal discharge syndrome. The diagnosis for the syndromes was according to the WHO flow chart for diagnosing sexually transmitted infections based on syndromic approach. Thirty-two (82.05%) patients acquired the disease for the first time and the rest 7 (17.95%) patients had recurrent disease. Only 5 (12.82%) patients were treated with their partners and the rest 34 (87.18%) patients were treated without their partners. The overall prevalence of sexually transmitted infections based on the syndromic approach was 10.4% (Table 3).

**Table 3.** Frequency distribution of clinical profiles of STI based on syndromic approach among HIV positive patients linked to ART clinic at AHMC.

| STI syndromes | Sex |  | ART started |  | Frequency (%) |
| --- | --- | --- | --- | --- | --- |
|  | Male (%) | Female (%) | Yes (%) | No (%) |  |
| Urethral discharge | 8(20.5%) | — | 7(87.5) | 1(12.5) | 8 (20.5%) |
| Genital ulcer | 3(7.69%) | 8(20.5%) | 11(100) | 0 | 11 (28.2%) |
| Vaginal discharge | — | 13(33.3%) | 12(92.3) | 1(7.7) | 13(33.3) |
| Scrotal swelling | 2 (5.12%) | — | 2(100) | 0 | 2 (5.12%) |
| Lower abdominal pain | — | 5(12.8%) | 5(100) | 0 | 5 (12.8%) |
| Inguinal bubo | — | — | — | — | — |
| Total | 13(33.34%) | 26(66.66%) | 37(94.8%) | 2(5.12%) | 39(100%) |

### WHO clinical staging of study subjects

Concerning WHO clinical stage, 2 (5.12%), 1 (2.56%), 1 (2.56%), and 4 (10.25%) of urethral discharge cases were in WHO stage I, II, III, and IV respectively. Four (10.25%), 2 (5.12%), 2 (5.12%) and 3 (7.69%) of genital ulcer cases were in WHO stage I, II, III, and IV respectively. Three (33.3%), 2 (2.56%), 3 (7.69), and 5 (12.82%) of vaginal discharge cases were in WHO stage I, II, III, and IV respectively. Two (2.56%), 1 (2.56) and (2.56%), lower abdominal pain cases were in WHO stage I, III, and IV respectively. Other STIS cases were scrotal swelling (Table 4).

**Table 4.** Distribution of WHO clinical staging and common STI/syndromic identified among HIV-positive patients linked to ART clinic at AHMC.

| Types of STI | WHO clinical staging of HIV/AIDS patients |  |  |  | Total No |
| --- | --- | --- | --- | --- | --- |
|  | Stage I<br>N (%) | Stage II<br>N (%) | Stage III<br>N (%) | Stage IV<br>N (%) |  |
| Urethral Discharge | 2(5.12) | 1(2.56) | 1(2.56) | 4(10.25) | 8(20.49) |
| Genital ulcer | 4(10.25) | 2(5.12) | 2(5.12) | 3(7.69) | 11(28.18) |
| Vaginal Discharge | 3(7.69) | 2(5.12) | 3(7.69) | 5(12.82) | 13(33.3) |
| Lower abdominal pain | 2(5.12) | — | 1(2.56) | 2(5.12) | 5(12.82) |
| Scrotal swelling | — | — | 1(2.56) | 1(2.56) | 2(5.12) |
| Inguinal bubo | — | — | — | — | — |
| Total no (%) | 11(28.18) | 5(12.82) | 8(20.49) | 15(38.42) | 39(100%) |

### Associated factors

After adjusting for all variables that were significant in bivariable analysis, the multivariable analysis revealed that age (<35), having multiple sexual partners and CD4 <500 have a significant statistical association with STI having a p-value of<0.05. In this study, the bivariable analysis result revealed that age (lower age group), having multiple sexual partners, alcohol, not using condoms regularly, previous STI, and lower CD4 count have significant statistical association with STI. Other variables like a new sexual partner and being unmarried have a p-value of 0.895 and 0.379 respectively with the bivariable analysis thus were not included in the multivariable analysis (Table 5).

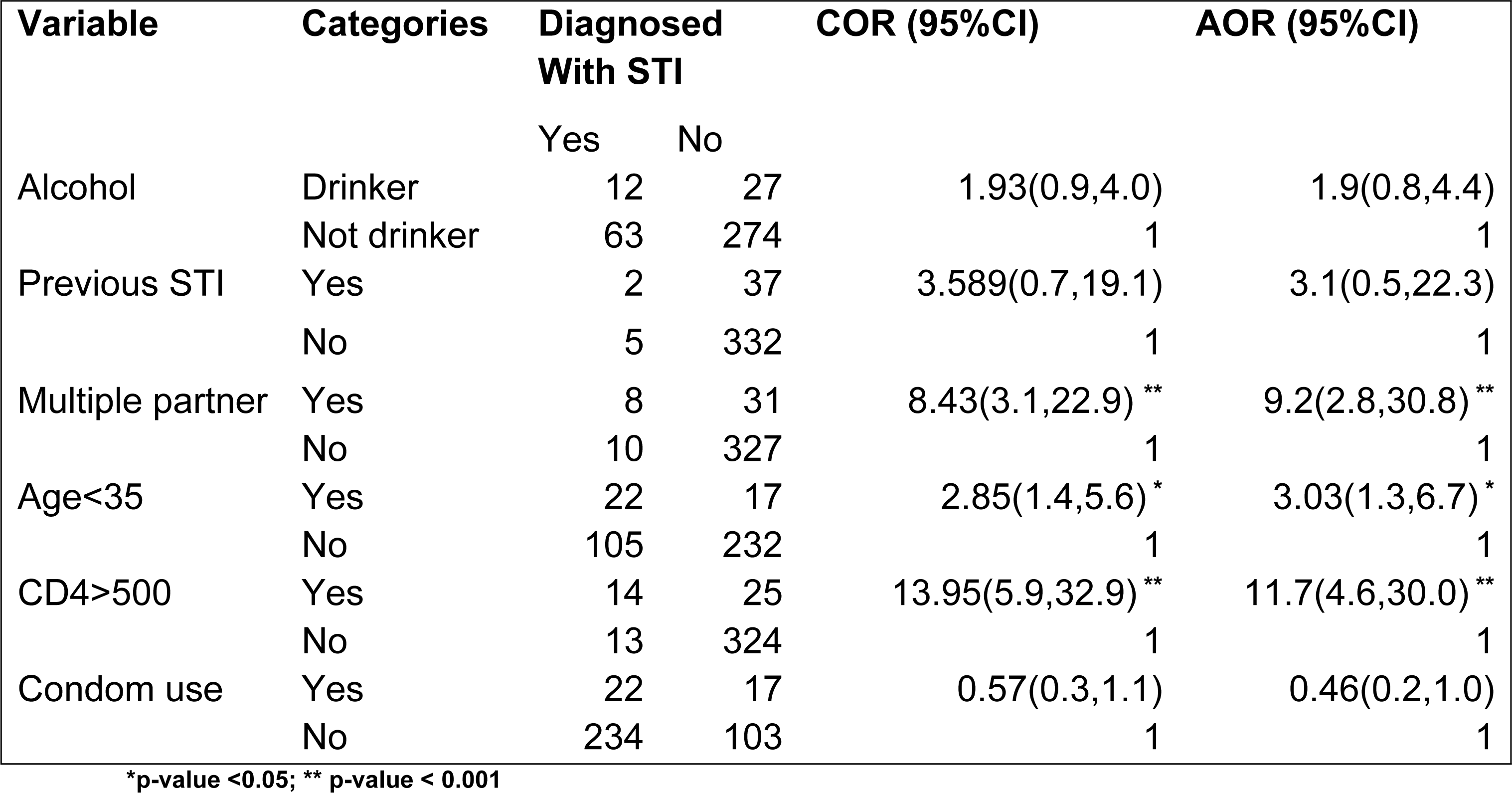

| Variable | Categories | Diagnosed With STI |  | COR (95%CI) | AOR (95%CI) |
| --- | --- | --- | --- | --- | --- |
|  |  | Yes | No |  |  |
| Alcohol | Drinker | 12 | 27 | 1.93(0.9,4.0) | 1.9(0.8,4.4) |
|  | Not drinker | 63 | 274 | 1 | 1 |
| Previous STI | Yes | 2 | 37 | 3.589(0.7,19.1) | 3.1(0.5,22.3) |
|  | No | 5 | 332 | 1 | 1 |
| Multiple partner | Yes | 8 | 31 | 8.43(3.1,22.9) ** | 9.2(2.8,30.8) ** |
|  | No | 10 | 327 | 1 | 1 |
| Age<35 | Yes | 22 | 17 | 2.85(1.4,5.6) * | 3.03(1.3,6.7) * |
|  | No | 105 | 232 | 1 | 1 |
| CD4>500 | Yes | 14 | 25 | 13.95(5.9,32.9) ** | 11.7(4.6,30.0) ** |
|  | No | 13 | 324 | 1 | 1 |
| Condom use | Yes | 22 | 17 | 0.57(0.3,1.1) | 0.46(0.2,1.0) |
|  | No | 234 | 103 | 1 | 1 |

\*p-value <0.05; \*\* p-value < 0.001

## DISCUSSION

This study revealed that among 376 HIV-positive patients 39(10.37%) have STI of them 8 patients (20.5%) had urethral discharge syndrome, 11 patients (28.2%) had genital ulcer syndrome, 2 patients (5.12) had the scrotal swelling syndrome,5 patients(12.8%) had lower abdominal pain syndrome, no inguinal bubo and 13 patients (33.3%) had vaginal discharge syndrome. And the result of this study is relatively low as compared to studies done in Peru, 27% and Thailand 47.0% [13, 25]. According to a study done in the town of Gondar, the prevalence was 383/1071 (35.8%) overall and 38.38% for vaginal discharge, 13.58% for urethral discharge, 2.87% for LAP, 2.09% for genital ulcers, and 1.04% for genital edema [24].

The relatively low prevalence of STI in this study can be explained by the small sample size, time of research (individual patients’ awareness of STI preventive strategies may have altered by the time of the study), and community-wide poor health-seeking behavior could all contribute to this low prevalence. One of the factors may be women’s ability to discern between abnormal and typical vaginal secretions. On the other hand, the magnitude of STIs was higher than the study conducted in Ethiopia at Adami Tulu woreda, which was (3.3%) [27]. This could be because, unlike the current investigation, the study in Adami Tulu woreda was community-based and did not focus on high-risk groups [7]. The result is comparable to results found in a study on Prevalence and associated factors of sexually transmitted infections among patients in the ART clinic in Mekele Ayder Hospital which was 30/353(8.5%) [21].

This study revealed that women had STIs at a greater rate (66.66% vs. 33.34%), on average. The rationale may be due to the higher proportion of females than males in this study as well as the fact that women are biologically more prone to STIs and are two to four times more likely than men to get STIs during unprotected vaginal intercourse. In this study, 17.94% (7/39) of STI patients had recurrent cases, 87.2% of STI patients did not bring their sexual partners for treatment, and 31.9% of STI patients did not regularly use condoms. When compared to the findings of a similar study done in Gondar town (no partner therapy 77%, recurring cases 2.1%), it was comparatively high. This could mean that the patients did not accept the advice and education they received from the health care provider at each subsequent visit, or that the health service providers (ART staff) did not fully counsel and convince the patients on the components of the STI syndrome management flow chart during each subsequent visit. Treatment failure brought on by several additional circumstances, such as the lack of a sexual partner, irregular condom use, antibiotic resistance, poor treatment compliance, HIV co-infection itself, and other opportunistic infections, could be the cause of the recurrence.

The finding of current study showed that 30 respondents (7.97%) had a new sexual partner within the last three months, of them 20 respondents (66.70%) had one new sexual partner, and the rest 33.3% had more than one new sexual partners and 68.1% of the participants used condom regularly during sexual intercourse. The results were low when compared to similar studies conducted in Ayder Hospital which revealed thirty-one respondents(8.8%) had a new sexual partner within the last three months, of them 18 respondents (58.1%) had one new sexual partner, and the rest 41.9% had more than one new sexual partners and 68.3% of the participants used condom regularly during sexual intercourse [21].

A study conducted in, Gondar, Ethiopia on Risky Sexual Practice and Associated Factors Among HIV Positive Adults showed that the study participants who were between the ages of 18 and 29 years engaged in hazardous sexual behavior 2.6 times (aOR = 2.63, 95% CI: 1.55, 4.47) more frequently than those who were 40 years of age or older [22]. and this was comparable to the results found in this study which revealed those under 35 years had 3.03 times more STI (aOR=3.03,95%CI;1.3,6.66) than those above 35 years. The reasons why the younger HIV patients were at higher risk of getting STIs than the elder ones might be that; they are a sexually active group and their awareness of safer sexual behavior might be lower. Single HIV patients were at higher risk of getting STIs than married HIV patients due to, having multiple sexual partners, being relatively younger (sexually active), high chance of having sex with commercial sex workers. The same study done in Gondar revealed Study participants with CD4 counts greater than 500/mm^3^ were 2.6 times (aOR = 2.58, 95% CI: 1.57, 4.24) more likely to engage in risky sexual behavior than participants with CD4 counts less than 350/mm^3^ [22]. While our study reveals that those with CD4 of <500 had 11.7 times(aOR-=11.7,95%CI;4.6,30) more STI than those with CD4 of > 500 the variation can be explained by the study done in Gondar tried to assess the sexual behaviors of patients with high CD4 count and low CD4 count and get a result that showed those with CD4 >500 had risky sexual behavior but that doesn’t always mean they end up with STI and this study shows those with CD4<500 had more STI than those >500 and it can be explained by depressed immunity.

Having multiple sexual partners is the third variable in this study that has a significant statistical association with STI. The study revealed that those with multiple sexual partners have 9.2 times (aOR = 9.2, 95% CI; 2.76–30.8) more STIs than those without multiple sexual partners. The result of this study shows a higher prevalence of STIs among those who have multiple partners compared to a study in Ethiopia, which revealed that those with multiple sexual partners have 2.29 times (aOR = 2.29, 95% CI: 1.05, 5.01) more STI than those without [26]. The above variation can be explained by the fact that the study only includes males who by different mechanisms are less prone to acquire sexually transmitted infections than females regardless of several sexual partners. However, our study included both males and females thus the rise in the association between STI and multiple sexual partners could be attributed to the female gender. It can also be explained by the study population in our study were HIV-positive people whose immunity is lower than that of the general population and having additional risk factors further increases the risk of acquiring STI.

### Limitation

Although the study come-up with crucial findings, it has some limitations. The cross-sectional nature of the study design does not allow causality ascertainment. Lack of recent studies conducted in Ethiopia with the same objective for comparison. Sexual issues are sensitive and the study subjects might not give information freely.

## Conclusion

The study revealed among 376 HIV-positive patients 39(10.37%) have STI and 337 (89.63%) have no STI. The magnitude of STIs was higher among females than males. Vaginal discharge, genital ulcer, and urethral discharge were more commonly reported STIs syndrome in this study. Age (lower age group), having multiple sexual partners, Alcohol, not using condoms, and lower CD4 count were the factors significantly associated with STI.

## Authors’ contributions

MS and AN were involved in conceiving the idea, study design, data analysis, and interpretation, writing the original manuscript. AN, MS and SN were involved the data handling and processing, data analysis, revising the manuscript and managing the overall progress of the study. The final manuscript was read and approved by all participated authors.

## Availability of data and materials

The datasets used and/or analyzed during the current study are available from the author (MS) upon reasonable request.

## Ethics approval

Ethical approval letter (Ref.No AHMC/636/21) was obtained from the ethical review committee of AHMC. A permission letter was written to the ART ward and the study was commencing after receiving formal permission from them. The Institutional Review Board approved the verbal consent. Data collectors-maintained confidentiality by excluding names or any other personal identifiers from data collection sheets and reports

## Consent for publication

Not applicable

## Data Availability

All relevant data are within the manuscript and its Supporting Information files.

## Acknowledgment

We would like to extend our deepest heartfelt gratitude to all study participants for their voluntary participation, and patience and for providing essential information. We are also grateful to data collectors and supervisors for their responsible data collection during the data collection period. Lastly, we like to thank Adama Hospital Medical College administration and health professionals for their cooperation and support during data collection.

## Acronyms/Abbreviations

AIDS: Acquired Immunodeficiency Syndrome
AHMC: Adama Hospital Medical College
ART: Antiretroviral Treatment; CD4– Cluster for Differentiation
CDC: Centers for Disease Control
HAART: Highly Active Antiretroviral Therapy
HIV: Human Immunodeficiency Virus
HTLV: Human Thymus Lymphocyte Virus
MOH: Ministry of Health
STI: Sexually Transmitted Infection
STD: Sexually Transmitted Disease
UNAIDS: United Nations Program on HIV/AIDS
WHO: World Health Organization

